# Drug Overdose Mortality Rates by Educational Attainment and Sex for Adults Aged 25–64 in the United States Before and During the COVID-19 Pandemic, 2015–2021

**DOI:** 10.1101/2023.05.19.23290239

**Authors:** Jay J. Xu, Marissa J. Seamans, Joseph R. Friedman

## Abstract

**Introduction:** Dramatic increases in U.S. drug overdose deaths involving synthetic opioids, especially fentanyl, beginning around 2014 have driven a marked progression in overall drug overdose deaths in the U.S., sharply rising after the onset of the COVID-19 pandemic. Disparities in drug overdose deaths by educational attainment (EA) during the fentanyl era of the drug overdose epidemic and its intersection with the COVID-19 pandemic have not been widely scrutinized.

**Methods:** Utilizing restricted-use mortality data from the National Vital Statistics System and population estimates from the American Community Survey, we estimated annual national age-adjusted mortality rates (AAMRs) from drug overdoses jointly stratified by EA and sex for adults aged 25–64 from 2015 to 2021. State-level AAMRs in 2015 and 2021 were also estimated to examine the geographic variation in the cumulative evolution of EA-related disparities over the course of the analysis period.

**Results:** Nationally, AAMRs rose fastest among persons with at most a high school-level education, whereas little to no change was observed for bachelor’s degree holders, widening pre-existing disparities. During the analysis period, the difference in national AAMRs between persons with at most a high school-level education and bachelor’s degree holders increased from less than 8-fold (2015) to approximately 13-fold (2021). The national widening of EA-related disparities accelerated amid the COVID-19 pandemic, and they widened in nearly every state. Among non-bachelor’s degree holders, national AAMRs increased markedly faster for males.

**Conclusions:** The widening disparities in drug overdose deaths by EA are a likely indicator of a rapidly increasing socioeconomic divide in drug overdose mortality more broadly. Policy strategies should address upstream socioeconomic drivers of drug use and overdose, particularly among males.

## Introduction

The illicit drug supply in the United States (U.S.) has become increasingly contaminated with clandestinely manufactured fentanyl and other ultra-potent synthetic opioids [1]. Dramatic increases in synthetic opioid-involved drug overdose deaths beginning around 2014 have driven a marked progression in overall drug overdose deaths in the U.S. [2], during what has been characterized as the “third wave” of the U.S. opioid overdose crisis [1]. Fatal drug overdoses sharply rose to unprecedented levels after the onset of the COVID-19 pandemic in March 2020, which occurred against the backdrop of increased substance use and an elevated prevalence of adverse mental health conditions relative to pre-pandemic levels [3–6]. Calendar year drug overdose deaths increased by an unprecedented 30% from 2019 to 2020 and topped 100,000 for the first time ever in 2021.

Nationally, this fentanyl-fueled surge in drug overdose deaths has most disproportionately affected non-Hispanic Black and American Indian and Alaska Native persons, especially among males [7], raising important public health concerns. During the COVID-19 pandemic, disparities in drug overdose deaths by race/ethnicity have received considerable research attention [8–16]. Here, we consider an alternative but interrelated disparities perspective on drug overdose deaths during the fentanyl era of the drug overdose epidemic, investigating differential drug overdose mortality by educational attainment (EA)–an important and modifiable social determinant of health [17] that has been found to be an independent risk factor for opioid overdose mortality [18]–and sex. Specifically, we estimate annual age-adjusted mortality rates (AAMRs) jointly stratified by EA and sex for U.S. adults aged 25–64 during the seven year period from 2015 to 2021, the last two years of which coincide with the COVID-19 pandemic. Furthermore, to examine geographic trends in the cumulative evolution of EA-related disparities in drug overdose deaths over the course of the analysis period, we estimate and contrast sex-specific AAMRs between adults aged 25–64 with at least some college education and adults aged 25–64 with at most a high school (HS) diploma/GED credential for each of the 50 states and the District of Columbia (D.C.) in 2015 and 2021.

## Materials and methods

### Data sources

We obtained access to and used the National Vital Statistics System (NVSS) restricted-use “All Counties” multiple cause of death research files [19] to identify all drug overdose deaths among U.S. residents occurring between January 1, 2015 and December 31, 2021. Consistent with the Centers for Disease Control and Prevention (CDC) definition of drug overdose deaths, we define drug overdose deaths as corresponding to International Classification of Diseases, 10^th^ Revision (ICD-10) codes X40–44 (unintentional), X60–64 (suicide), X85 (homicide), or Y10–14 (undetermined intent) as the underlying cause of death [2]. We limit our attention to drug overdose deaths among adults aged 25–64 because the vast majority of eventual bachelor’s degree holders will have received their bachelor’s degree by age 25 and because EA is less reliably reported on U.S. death certificates for decedents aged 65+ [20]. Furthermore, this age range captures the vast majority of the at-risk population from the drug overdose epidemic, comprising over 86% of total drug overdose deaths between 2015 and 2021. Within this age range, we considered the following age subgroups for mortality rate calculations: 25–34, 35–44, 45–54, and 55–64. Mortality statistics jointly stratified by EA and sex reported in official NVSS publications also restrict attention to this age range and consider these age subgroups [21].

Within the U.S., death certificates are submitted by 52 jurisdictions to the NVSS: New York City, New York State (minus New York City), the remaining 49 states, and D.C. The U.S. Standard Certificate of Death (SCoD) was revised in 2003 [22–24] from its previous revision in 1989 [25], and part of the 2003 revision involved transitioning from a year-based categorization of EA to a degree-based categorization, which individual jurisdictions gradually adopted over time. For the present analysis, we categorized EA into three categories derived from the 2003 U.S. SCoD revision-based EA reporting standard: (i) HS/GED or Less, (ii) Some College Education (including obtaining an Associates degree), and (iii) Bachelor’s Degree or More.

Rhode Island began reporting EA on death certificates in mid-2015 [26], the last jurisdiction to begin doing so. Furthermore, 2018 was the first full calendar year in which all U.S. jurisdictions reported EA on death certificates submitted to the NVSS according to the 2003 U.S. SCoD revision-based EA reporting standards; this is because in 2015, 2016, and part of 2017, EA reported on death certificates from West Virginia still used the year-based categorization according to the 1989 U.S. SCoD revision-based EA reporting standard [27], and in 2015, Alabama still used the 1989 U.S. SCoD revision-based EA reporting standard [26] before making the transition in 2016 [28]. From 2015 to 2021, there were 58 drug overdose deaths with unreported EA and age, 19 drug overdose deaths with reported EA but unreported age, and 13,701 drug overdose deaths aged 25–64 with reported age but unreported EA. Of the 452,706 drug overdose deaths from 2015 to 2021 with reported age in the 25—64 age range, 3.0% had unreported EA.

State population estimates (as well as population estimates for D.C.) within EA-sex-age strata for years 2015–2019 and 2021 were obtained from American Community Survey (ACS) 1-year estimates. The COVID-19 pandemic substantially disrupted ACS survey operations in 2020, affecting its quality and delaying its annual data release. The cumulative effect of the COVID-19 pandemic on 2020 ACS operations included obtaining a respondent sample whose EA distribution was excessively upwards-skewed [29]. As a result, the U.S. Census Bureau decided that for the 2020 ACS, it would not release its standard suite of 1-year data products, releasing instead a limited collection of experimental population estimates, which did not include state population estimates within EA-sex-age strata [30, 31]. As such, for our analysis, we opted to linearly interpolate state population sizes in 2020 within each EA-sex-age stratum by fitting state-EA-sex-age stratum-specific univariate linear regression models of annual population estimates as a function of year to years 2015–2019 and 2021 and obtaining fitted population estimates for 2020.

### Statistical Analysis

For the 2,702 drug overdose deaths occurring in West Virginia and Alabama between 2015 and 2017 with EA reported according to the 1989 SCoD revision-based EA reporting standard, we deterministically imputed their (trichotomized) EA categories using the following procedure: (i) decedents recorded as having completed at most 12^th^ grade were classified as HS/GED or Less, (ii) decedents recorded as having completed 1–3 years of college were classified as Some College Education, and (iii) decedents recorded as having completed 4 or more years of college were classified as Bachelor’s Degree or More. Next, consistent with a missing at random assumption [32], we deterministically imputed the unknown EA categories of the 13,701 drug overdose deaths with unreported EA but reported age according to the following procedure: within each year-jurisdiction-sex-age stratum, we applied the relative proportions of the EA categories for drug overdose deaths with reported EA and age to the drug overdose deaths with unreported EA but reported age, rounding as necessary. For simplicity, we omitted the 77 drug overdose deaths with unreported age from all mortality rate calculations.

Following the imputation processes and the removal of drug overdose deaths with unreported age, we calculated annual national AAMRs stratified by EA and sex from 2015 to 2021. AAMRs were calculated using direct age adjustment [33], standardized to the 2000 U.S. Standard Population [34, 35]. Furthermore, to examine the geographic variation in the cumulative evolution of EA-related disparities over the course of the analysis period, we combined the top two EA categories and calculated AAMRs in 2015 and 2021 for each of the 50 states and D.C., jointly stratified by EA and sex. Then, for each year-location-sex stratum, we calculated the AAMR ratio between adults aged 25–64 with at most a HS-level education and adults aged 25–64 with at least some college education as a measure of the disparity in drug overdose deaths by EA. We combined the top two EA categories due to small drug overdose death counts among bachelor’s degree holders in many states.

The entire analysis was performed using R version 4.2.1 [36]. This study was deemed exempt from review by the University of California, Los Angeles Institutional Review Board.

## Results

Figure 1 illustrates the estimated annual AAMRs and their annual changes stratified by EA and sex from 2015 to 2021. Table 1 in the Appendix presents these AAMRs as well as the underlying age-specific mortality rates (ASMRs). For both males and females during the analysis period, AAMRs increased most rapidly among those with at most a HS-level education, and little to no temporal variation in AAMRs was observed for bachelor’s degree holders relative to the lower two EA categories, substantially widening pre-existing disparities in drug overdose deaths by EA. Among males with at most a HS-level education, the AAMR was over 2.5 times higher at the end of the analysis period than at the beginning of the analysis period, increasing from 59.5 to 149.3 deaths per 100,000, while the corresponding AAMR for females also more than doubled from 36.3 to 72.9 deaths per 100,000. Among males with some college education, the AAMR increased more than two-fold from the beginning to the end of the analysis period, from 35.5 to 74.9 deaths per 100,000, while the corresponding AAMR for females rose from 25.6 to 43.7 deaths per 100,000. The annual AAMR trajectories for males with some college education and females with at most a HS-level education were very similar during the analysis period. The AAMR for male bachelor’s degree holders ticked up marginally from the beginning to the end of the analysis period from 7.5 to 11.5 deaths per 100,000, and the AAMR for female bachelor’s degree holders remained virtually unchanged over the course of the analysis period, equal to 5.0 deaths per 100,000 in 2015 and 5.6 deaths per 100,000 in 2021. Notably, from 2017 to 2018, AAMRs actually decreased within each EA-sex stratum, albeit marginally. For both male and female non-bachelor’s degree holders, AAMRs grew most rapidly during the first two years coinciding with the COVID-19 pandemic.

**Table 1:**
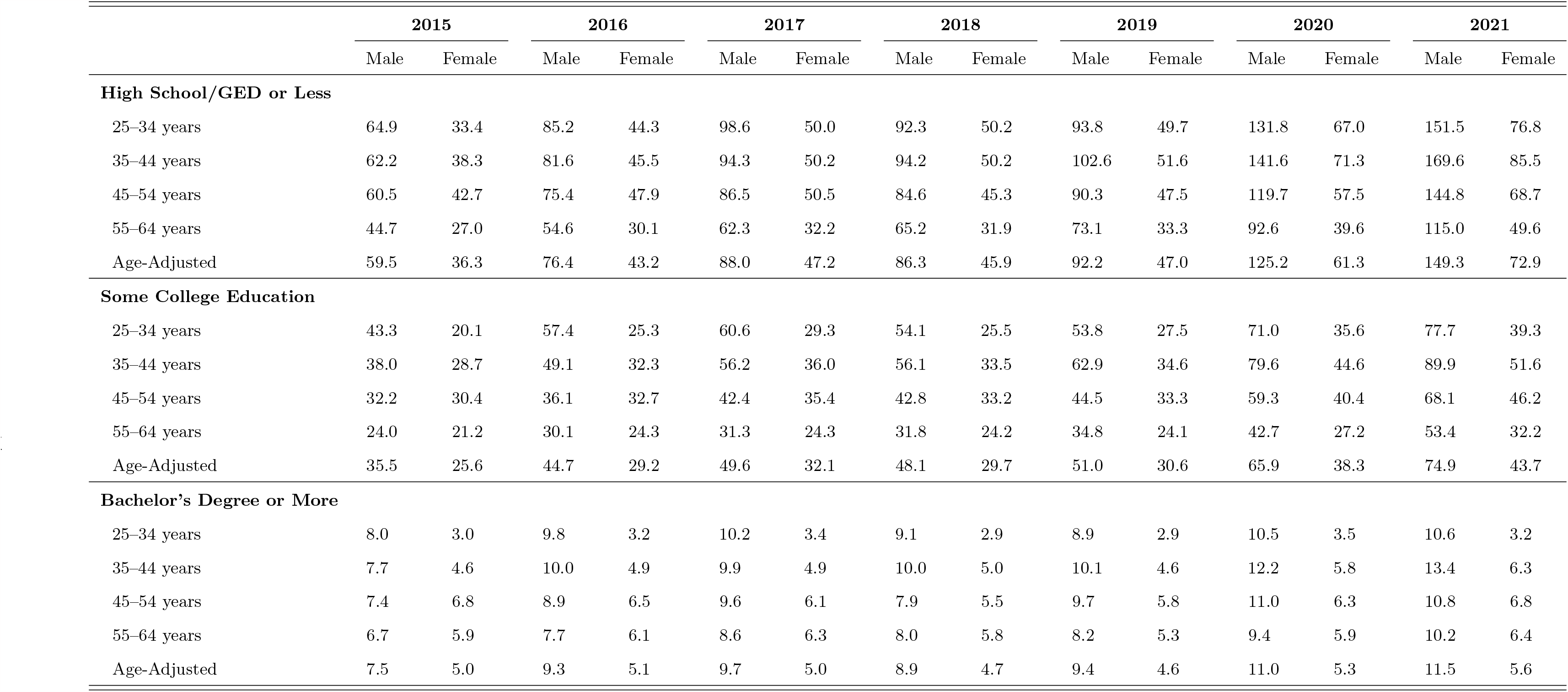
Estimated annual age-specific and age-adjusted drug overdose mortality rates per 100,000 by educational attainment and sex for U.S. adults aged 25–64, 2015–2021.

**Figure 1:**
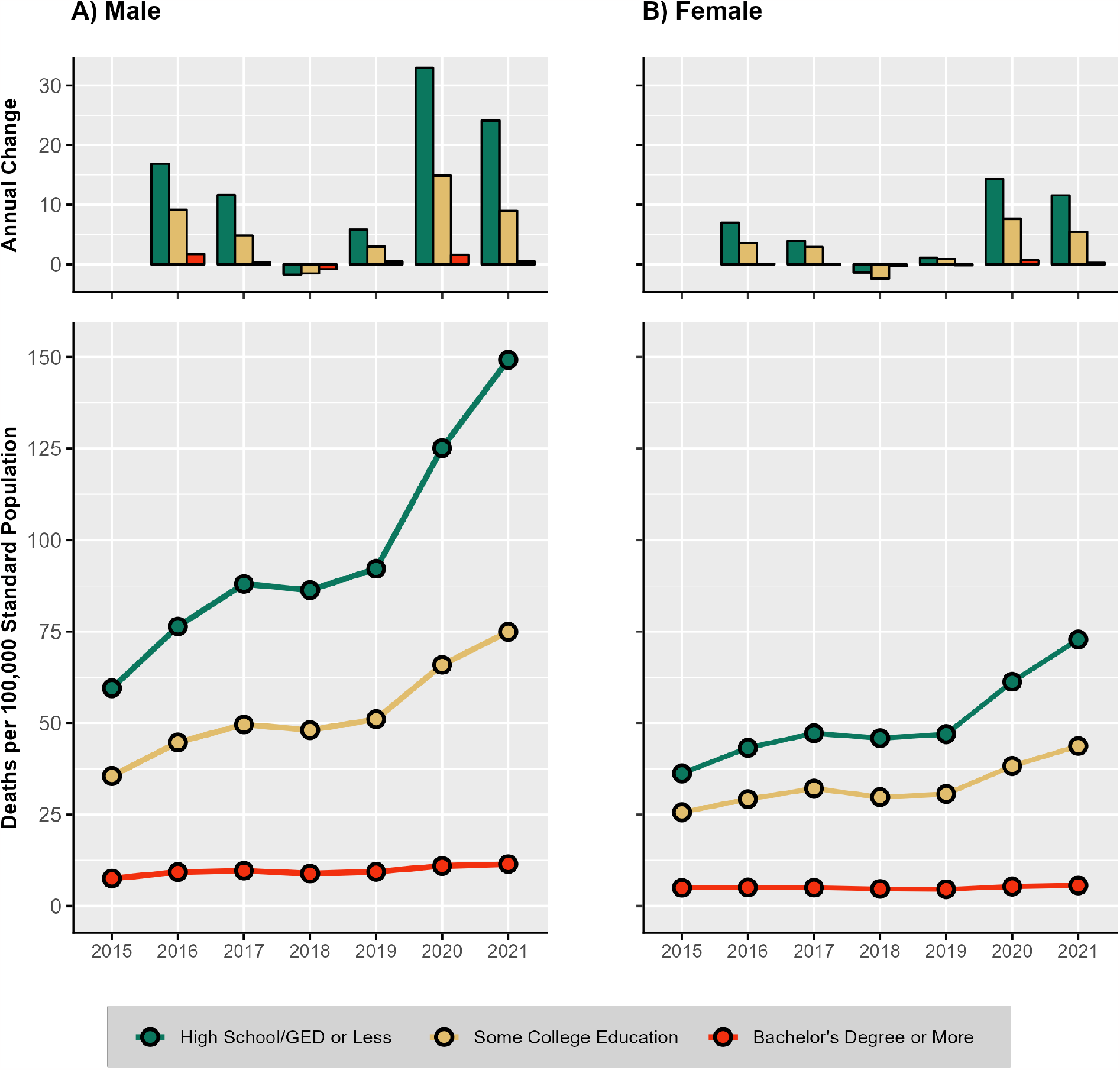
Estimated annual age-adjusted drug overdose mortality rates and their annual changes per 100,000, jointly stratified by educational attainment and sex for U.S. adults aged 25–64, 2015 to 2021.

In 2015, the AAMR for males with at most a HS-level education was nearly 8 times that of male bachelor’s degree holders, and the AAMR for females with at most a HS-level education was over 7 times that of female bachelor’s degree holders. By 2021, these disparities widened to a greater than 13-fold difference among males and a nearly 13-fold difference among females. On the absolute scale, male AAMRs increased more rapidly than female AAMRs within each EA category during the analysis period, with the differential rate of growth inversely associated with EA, widening pre-existing sex disparities. The sex disparity in AAMRs within each EA category widened most rapidly during the first two years coinciding with the COVID-19 pandemic. By 2021, the male AAMR became over twice as high as the female AAMR within the lowest and highest EA categories.

With the exception of female bachelor’s degree holders aged 45–54, mortality rates for each EA-sex-age stratum increased from 2015 to 2021, and with the exception of female bachelor’s degree holders, the absolute increases in ASMRs from 2015 to 2021 were highest in the 35–44 age group within each EA-sex stratum. For all EA categories, male ASMRs were highest in the 25–34 age group in 2015 but became highest in the 35–44 age group in 2021. Female ASMRs were highest in the 45–54 age group for all EA categories in 2015, but in 2021, they became highest in the 35–44 age group for non-bachelor’s degree holders and remained highest in the 45–54 age group for bachelor’s degree holders.

Figure 2 illustrates the ratio of the AAMRs between adults with at least some college education and those with at most a high school diploma/GED credential in 2015 and 2021 for each location-sex stratum. The 2015 and 2021 ratios for the U.S. overall are included for reference. AAMR ratios were greater than 1.0 for each of the 204 considered year-location-sex strata, and in 94 out of the 102 (92.2%) location-sex strata, the 2021 AAMR ratio exceeded the 2015 AAMR ratio. Therefore, while there was substantial variation in the magnitude of EA-related disparities in drug overdose deaths across states at both the beginning and the end of the analysis period, evidenced by the substantial variability in the AAMR ratios, the widening of disparities in drug overdose deaths by EA during the analysis period at the national level was the result of a near-universal widening of these disparities at the state level.

**Figure 2:**
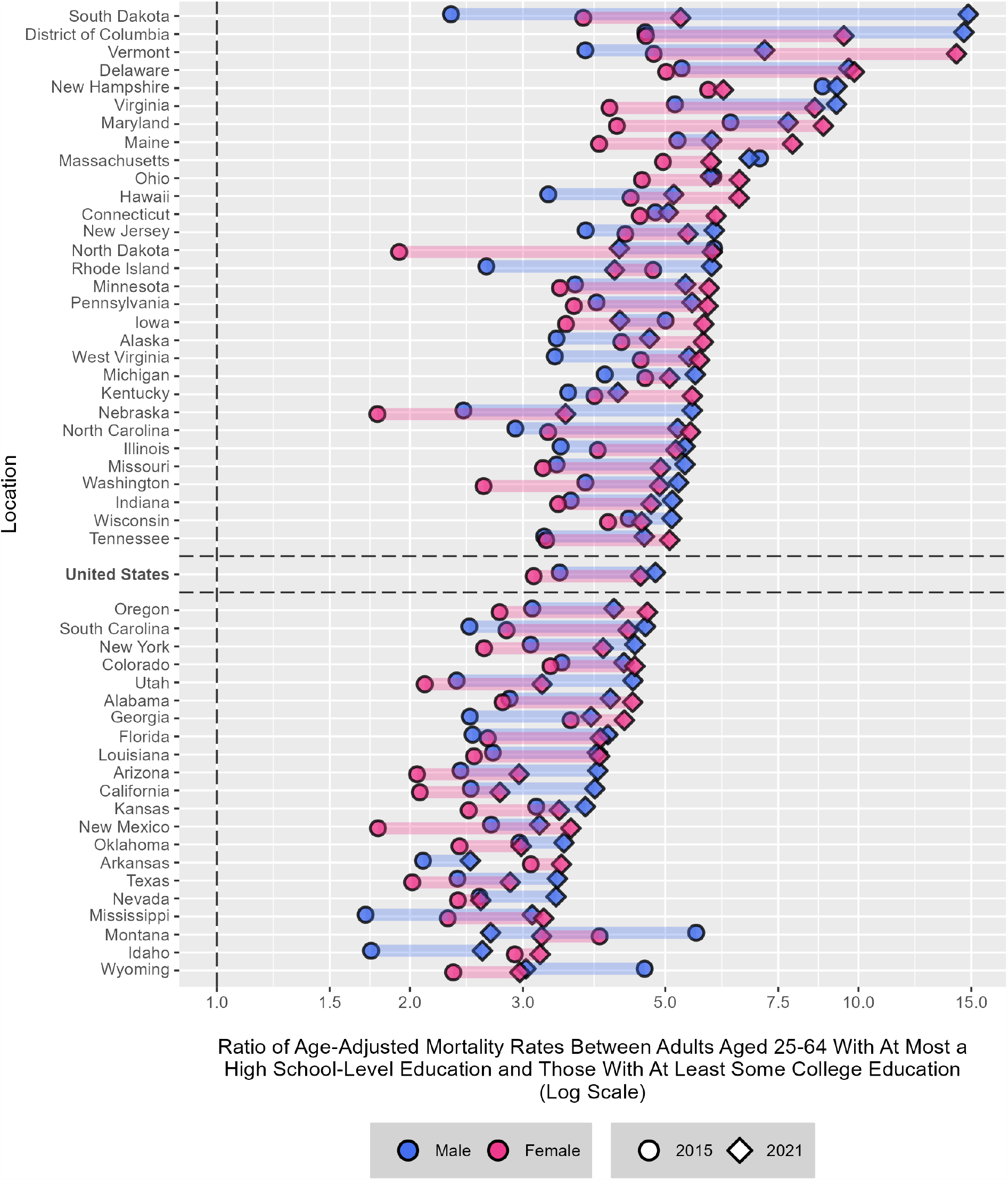
Estimated ratios of age-adjusted drug overdose mortality rates between U.S. adults aged 25–64 with at least some college education and those without in the 50 states and D.C., stratified by sex.

## Discussion

In this study, we documented empirical evidence that the ongoing drug overdose epidemic, which has been rapidly intensified by the proliferation of illicitly manufactured synthetic opioids, especially fentanyl, poses disproportionate risks to populations of lower EA, especially among males. EA-related disparities in fatal drug overdoses gradually worsened in years prior to the COVID-19 pandemic, a trend that was exacerbated during the COVID-19 pandemic. Viewing the drug overdose epidemic from the vantage point of heterogeneous risk profiles between population subgroups with different levels of EA, and how it intersects with sex, offers valuable insights into which segments of the U.S. population are at greatest and increasing risk. Given strong correlations between EA and a plethora of social and economic outcomes [37, 38], our study findings indicate more broadly that persons who use drugs (PWUD) of lower socioeconomic status (SES) are at increased and increasing vulnerability to the increasingly toxic illicit drug supply.

During the COVID-19 pandemic, the combination of marked shifts in the composition of the U.S. illicit drug supply, reduced access to drug addiction treatment services, increased psychosocial and financial stress, and higher levels of using drugs alone likely contributed to both the massive surge in drug overdose deaths and the accelerated widening of the disparities in drug overdose deaths by EA – and by SES more broadly. In response to U.S. travel restrictions and border closures enacted during the COVID-19 pandemic that made it harder to smuggle illicit drugs into the U.S. by land, drug cartels reportedly quickly adapted to their new operating environment, focusing more on the manufacturing of fentanyl-laced counterfeit prescription pills that are easier to conceal and transport into the U.S. by postal mail [39–41]. The number of counterfeit prescription pills containing fentanyl seized by law enforcement soared after the onset of the COVID-19 pandemic [42], and biospecimen drug testing studies documented increased fentanyl consumption after the onset of the COVID-19 pandemic in 2020 [43, 44], substantiating public health fears that U.S. illicit drug markets became even further permeated by fentanyl during the COVID-19 pandemic.

Disruptions to drug addiction treatment services during the COVID-19 pandemic compounded pre-existing access challenges to drug addiction treatment services for lower-SES PWUD, likely contributing to greater illicit drug procurement and use. The economic fallout precipitated by the COVID-19 public health crisis undoubtedly impacted lower-SES persons disproportionately, who were likely more susceptible to begin or more frequently use illicit drugs to cope with stress arising from COVID-19 pandemic-attributable financial hardship and the ensuing symptoms of anxiety and depression [45]. Increased social isolation during the COVID-19 pandemic as a result of government stay-at-home orders and physical distancing likely led to an increased prevalence of solitary drug use [46, 47], especially among lower-SES persons concurrently experiencing financial hardship arising from the COVID-19 pandemic, incurring the added risk of overdosing without other persons available to administer potentially life-saving measures such as naloxone.

Nationally, the male AAMR (all ages) has been consistently higher than the female AAMR throughout the 21^st^ century [48, 2], but a pronounced widening of the sex gap began around 2015, which markedly accelerated during the first two years coinciding with the COVID-19 pandemic [2]. Our study demonstrates that substantial increases in drug overdose deaths among males aged 25–64 with at most a HS-level education were the driving force behind the pronounced widening of the sex gap in drug overdose mortality between 2015 and 2021; strikingly, we estimate that during each of the first two years coinciding with the COVID-19 pandemic, males with at most a HS-level education comprised over half of all of the drug overdose deaths that occurred among adults aged 25–64. Further investigation into the underlying supply- and demand-side factors behind why, since around 2015, the national drug overdose mortality burden among adults aged 25–64 has become increasingly disproportionately borne by males of lower EA at such a rapid pace is warranted.

Our study is subject to a number of limitations. First, our study focuses exclusively on the 25– 64 age range, so our study findings may not reflect trends in the population aged 65+, which has also seen steep increases in AAMRs during the fentanyl era [49]. Second, EA on death certificates is subject to misreporting [20]. Third, our procedure to deterministically impute the unknown EA categories of the examined drug overdose deaths with unreported EA but reported age assumed that the distribution of EA is independent of reporting status within year-jurisdiction-sex-age strata, an assumption that is unverifiable. Fourth, our procedure to deterministically impute the EA categories of drug overdose deaths occurring in Alabama and West Virginia that reported EA according to the 1989 SCoD revision-based EA reporting standard is subject to error, although the EA categories for the subset of these decedents reported as having completed at most 12^th^ grade were presumably all (or virtually all) correctly imputed as HS/GED or Less. Because both imputation tasks were performed determinisitically, we did not represent the imputation uncertainty in any of the ASMR or AAMR estimates. However, given the small percentage (3.0%) of the examined drug overdose deaths with imputed EA category values, doing so by utilizing a stochastic imputation procedure for each imputation task would not be expected to meaningfully alter the conclusions of our analysis.

## Conclusions

Our study findings demonstrate that U.S. policy makers must be cognizant of the already wide and widening disparities in drug overdose deaths by EA, a likely indicator of a rapidly increasing socioeconomic divide in drug overdose mortality more broadly, and that males experienced an increasingly disproportionate risk of drug overdose mortality over the course of the analysis period, especially those with lower levels of EA. Disparities aside, given the current unprecedented magnitude of drug overdose mortality in the U.S. and its upward trajectory, stemming the tide of fatal drug overdoses should be a top national public health priority. In the near term, expanding access to drug addiction treatment services, including pharmacotherapy [50, 51] and telehealth care [52], as well as lowering logistical and financial barriers to medications for OUD [53], represent critical evidence-based policies toward that end. Investments in modern overdose surveillance systems [54], ideally nationally-linked, publicly accessible, and updated in real time, are also warranted to help promptly identify areas of greatest need and guide resource allocation. Policy strategies should consider the unique needs of lower-SES PWUD, especially among males, and tailor interventions accordingly. Investments pertaining to upstream socioeconomic determinants of drug use, including access to preventative care, mental health services, and stable housing, are not only important for drug overdose prevention [55] but are also important for the prevention of other socially bound public health hazards.

## Data Availability

The mortality data used in the present study were obtained by submitting an application to the National Center for Health Statistics, available at https://www.cdc.gov/nchs/data/nvss/nchs-research-review-application.pdf. The U.S. state population estimates used in the present study were obtained from publicly available American Community Survey 1-year estimates, available from the U.S. Census Bureau Microdata Access Tool: https://data.census.gov/mdat/#/. The 2000 U.S. Standard Population is publicly available at https://seer.cancer.gov/stdpopulations/stdpop.singleages.html.

## Appendix

## Notes

### Competing Interest Statement

The authors have declared no competing interest.

### Funding Statement

This study did not receive any funding.

### Author Declarations

This study was deemed exempt from review by the University of California, Los Angeles Institutional Review Board.

### Summary of Updates

We added a reference and made a couple of minor edits.

